# Saliva as a testing sample for SARS-CoV-2 detection by RT-PCR in low prevalence community settings

**DOI:** 10.1101/2020.10.20.20216127

**Authors:** Didzis Gavars, Mikus Gavars, Dmitry Perminov, Janis Stasulans, Justine Stana, Zane Metla, Jana Pavare, Eriks Tauckels, Egils Gulbis, Uga Dumpis

## Abstract

**Objectives:** The number of COVID-19 cases is increasing globally and there is an urgency for a simple non-invasive method for the detection of SARS-CoV-2. Our study aimed to demonstrate that saliva can be used as a specimen for SARS-CoV-2 detection notably for the screening of extensive population groups via pooling.

**Methods:** To demonstrate that saliva is an appropriate specimen for SARS-CoV-2 detection a field study including 3,660 participants was performed between September 29 and October 1, 2020. We collected paired nasopharyngeal/oropharyngeal swabs (NPS) and saliva specimens and processed them within 24 hours of collection. We performed 36 serial measurements of 8 SARS-CoV-2 positive saliva samples to confirm the stability of the specimen and completed 37 pools of saliva samples by adding one positive specimen per pool.

**Results:** Saliva specimens were stable for testing for up to 24 hours. Overall, 44 salival samples (1.2%) tested positive for SARS-CoV-2 during the field study. The results of saliva samples were consistent with those obtained from NPS from the same patient with 90% sensitivity (95% CI 68.3%-98.7%) and 100% specificity during the first two weeks after the onset of symptoms. Using pooling strategy 796 RT-PCR tests were performed. All pools showed 100% positivity in different pooling proportions.

**Conclusions:** Our findings demonstrate that saliva is an appropriate specimen for pooling and SARS-CoV-2 screening with accurate diagnostic performance. Patient-performed simple specimen collection allows testing an extensive number of people rapidly, obtaining results of the spread of SARS-CoV-2 and allowing authorities to take timely measures.

## Introduction

Since the beginning of 2020 massive resources have been dedicated to control and mitigate the COVID-19 pandemic. Testing capacity and accessibility are crucial in monitoring COVID-19 outbreaks and allowing for the adjustment of measures put into place to reduce community transmission (1).

Viral nucleic acid detection using the real-time polymerase chain reaction (RT-PCR) assay remains the gold standard for the detection of SARS-CoV-2 and to diagnose COVID-19 (2). Due to the increased demand for sampling and testing materials globally considerable constraints remain to conduct widespread screening for SARS-CoV-2. It is especially challenging for counties with low prevalence levels as global suppliers allocate materials and decide delivery schedules based on the prevalence of the virus.

Shortages of supplies and medical staff, logistical hurdles, as well the unpleasantness associated with obtaining a RT-PCR specimen making it difficult to practice the procedure on children and causing irritation for those who need to undergo repeated tests, all these factors have naturally accelerated research into alternative specimens and sampling methods for COVID-19 testing (3,4).

On September 21, 2020, Latvia had a 14-day cumulative number of COVID-19 cases of 5.1 per 100,000 people. By October 10 that number had risen to 66. During this period, laboratories experienced a significant overload (5). Rapid and efficient solutions to increase testing capacity were needed.

The objective of this study was to confirm that saliva is a suitable specimen comparable to a nasopharyngeal/oropharyngeal swab for SARS-CoV-2 detection in specific populations. We also evaluated the pooling approach to employ it in field conditions for extensive screening of high-risk groups.

## Methods

### Patient sample collection

A single-center study was performed including all consented patients attending the laboratory for SARS-CoV-2 testing between May 12 and October 19, 2020. For the collection of the saliva samples, self-collection kits were distributed consisting of a specimen vial, registration form, alcohol pads for disinfection, and two safety plastic bags. Patients were requested to collect their saliva in an isolated location (keeping at least five meters distance from other persons or objects), preferably outdoors, at home, or in their car. The saliva was collected in a container without any additive and delivered to the laboratory within 24 hours. Paired nasopharyngeal/oropharyngeal swabs (NPS) for RT-PCR were collected using the standard method used in our laboratory (6).

### Sample preparation and analysis using RT-PCR

Immediately upon arrival at the laboratory, the samples were pretreated by adding 1 ml of phosphate-buffered saline (PBS). The extraction of RNA and RT-PCR testing was identical for both swab and pre-treated saliva samples. Viral RNA was extracted with standard commercial extraction methods (QIAGEN, Germany and LifeRiver, China) used in our laboratory. RT-PCR was performed using our laboratory-developed and validated test method which detects S and N genes of the SARS-CoV-2 virus. Our validated and verified limit of detection (LOD) was 1 cp/rxn.

## Results

### Stability of saliva specimens

To confirm the stability of saliva samples, we performed 36 serial tests on 8 primary SARS-CoV-2 positive saliva samples collected from patients 48 hours after their NPS tested positive. The data are summarized in Tables 1-3. Testing was randomized by different time intervals, the time of day, and the shifts of technicians. It includes repeated testing of two samples after 22h (Table 2) and serial repeats of one sample (Table 3) with Ct value reporting.

**Table 1.** Run No. 1 on saliva stability measurements of 5 samples collected 2-4h before the first testing

|  | **0** | **after 8h** | **after 24h** | **after 25h** |
| --- | --- | --- | --- | --- |
|  | *1st 14:00* | *2nd 22:00* | *3rd 14:00* | *4th 15:00* |
| 1 | Positive | Positive | Positive | Positive |
| 2 | Positive | Positive | Positive | Positive |
| 3 | Positive | W- Positive | Positive | W- Positive |
| 4 | Positive | Positive | Positive | No sample left |
| 5 | Positive | W- Positive | Positive | W- Positive |

Abbreviations: W-Positive - weak positive result, h - hours

**Table 2.** Run No. 2 on saliva stability serial measurements of two samples collected 2-4h before first testing

|  | **0** | **after 12h** | **after 22h** | **after 22h rep.** |
| --- | --- | --- | --- | --- |
|  | *1st 12:00* | *2nd 00:00* | *3rd 10:00* | *4th 10:00* |
| 6 | W- Positive | Positive | Positive | Positive |
| Ct | 35 | 36 | 37 | 37 |
| 7 | Positive | Positive | Positive | Positive |
| Ct | 32 | 33 | 30 | 31 |

Abbreviations: W-Positive - weak positive result, rep. - repeats, h – hours

**Table 3.** Run No. 3 on saliva repeatability measurements of one sample collected 2 - 4h before the first testing with indicated Ct values

|  | 0 | after 3h | after 12h | after 24h |
| --- | --- | --- | --- | --- |
|  | *1st 11:00* | *2nd 14:00* | *3rd 02:00* | *4th 11:00* |
| 8 | Positive | Positive | Positive | Positive |
| Ct | 34 | 30 | 33 | 35 |
| 8 rep. | Positive | Positive | Positive | Positive |
| Ct | 34 | 27 | 31 | 35 |

Abbreviations: rep. - repeats, h – hours

All results confirmed a positive result after 22-25h of the first test being conducted. Samples were stored refrigerated at 2-6°C between tests.

### Comparison of saliva and nasopharyngeal/oropharyngeal samples

Between May 13 and September 21, 2020, 431 tests on saliva samples were performed, and the results were compared with paired NPS RT-PCR test results. Using NPS RT-PCR as the reference, there were 104 PCR positive samples and 327 negative patients, including 125 pediatric (age 5-17 years (average 11.3 years)) samples. The observed RT-PCR positive results were divided into groups according to the number of days after the onset of symptoms: 0-14 days (22 patients); 15-30 days (52 patients); 31-70 days (20 patients) and 10 asymptomatic patients. The results of the comparison of the mentioned subgroups are presented in Table 4.

**Table 4.** Saliva samples compared to the results of nasopharyngeal/ oropharyngeal swabs

|  |  | NPS |  |  |  |  |  |  |  |
| --- | --- | --- | --- | --- | --- | --- | --- | --- | --- |
| Days after the start of symptoms |  | 0-14 |  | 15-30 |  | 31-70 |  | Asymptomatic |  |
| RT-PCR |  | Positive | Negative | Positive | Negative | Positive | Negative | Positive | Negative |
| Saliva | Positive | 18 | 0 | 22 | 3 | 5 | 2 | 5 | 1 |
|  | Negative | 2 | 2 | 12 | 15 | 2 | 11 | 2 | 2 |
| Sensitivity % |  | 90 |  | 64.7 |  | 71.4 |  | 71.4 |  |
| Specificity % |  | 100 |  | 83.3 |  | 84.6 |  | 66.7 |  |

Abbreviations: NPS - nasopharyngeal/ oropharyngeal swabs, RT-PCR – real-time polymerase chain reaction

Sensitivity and specificity for the respective groups were: for 0-14 days 90% (95%CI 68.3%-98.7%) and 100% (95%CI 22.4%-100%); for 15-30 days 64.7% (95% CI 46.5%-80.3%) and 83.3% (95%CI 58.9%-96.4%); for 31-70 days 71.4% (95%CI 29.0%-96.3%) and 84.6% (95%CI 54.6%-98.1%); for asymptomatic 71.4% (95%CI 29.0%-96.3%) and 66.7% (95%CI 0.8%-90.6%). The mean Ct values for the groups were 33.9 for NPS and 33.8 for saliva (0-14 days), 34.8 for NPS and 34 for saliva (15-30 days), and 36.5 for NPS and 39 for saliva in the asymptomatic patient group.

### Pooling of saliva

To evaluate the testing of pooled saliva samples, 37 pools were constituted. There were 15 pools of 5 samples (4 negatives + 1 positive), 13 pools of 10 samples (9 negatives + 1 positive), and 9 pools of 20 samples (19 negatives + 1 positive). The RT-PCR testing for each pool was repeated twice. The total number of measurements was 74. All results were 100% positive.

When using pooling, the number of tests needed per sample was calculated as follows: the total number of tests needed to test all samples is divided by the total number of samples. For example, if 500 samples are being tested in pools of 10, it means 50 tests are initially needed. With a positivity rate of 1%, a maximum of five pools will contain a positive specimen (if each positive sample is in a different pool). The 50 samples from these five pools will have to be retested individually (each of the five positive pools contains 10 samples). Therefore, a total of 100 tests are needed to evaluate all 500 samples and the number of tests per sample is 100/500 = 0.2.

The positivity rate in Latvia on September 23, 2020, was 0.5%.

The approximate positivity rate at which the pooling of 5 samples becomes more efficient than the pooling of 10 samples is 3% (2-4% depending on test price, laboratory load, tests per sample).

### Field study

To evaluate the convenience of saliva pool testing by RT-PCR in field conditions, the town of Kuldiga (total population 10,352) with an ongoing outbreak of COVID-19 in a textile factory was selected. In collaboration with local authorities, 4,100 saliva self-sampling kits were distributed to inhabitants at a specially established distribution point. There were four persons dedicated to the distribution of the kits and four drivers to deliver the samples by car to the testing laboratory in Riga (150 km one way). During the study period, 3,660 saliva samples were collected (response rate 91.5%), delivered to the laboratory, and tested in pools of 10 samples. There were a total of 366 pools. In the first round of testing 43 pools were found to be positive and samples from these pools were re-tested individually. The saliva samples from 44 patients were confirmed as positive by RT-PCR. Mean Ct values of pooled samples were 13% higher than individually tested Ct (31.6 versus 27.6). The positivity rate of the tested population was 1.2%. In total 796 RT-PCR tests were performed. At the time of the sample collection, 68.2% (30/44) of patients did not report any symptoms on the questionnaire form.

Nasopharyngeal swab tests and saliva tests were compared with the number of new COVID-19 cases. The findings illustrate that the second peak in new cases occurred 6-7 days after the initial peak. Such findings correspond with the mean incubation period of the SARS-CoV-2. The results reflect a decrease in new cases in the following observation period (Figure 1).

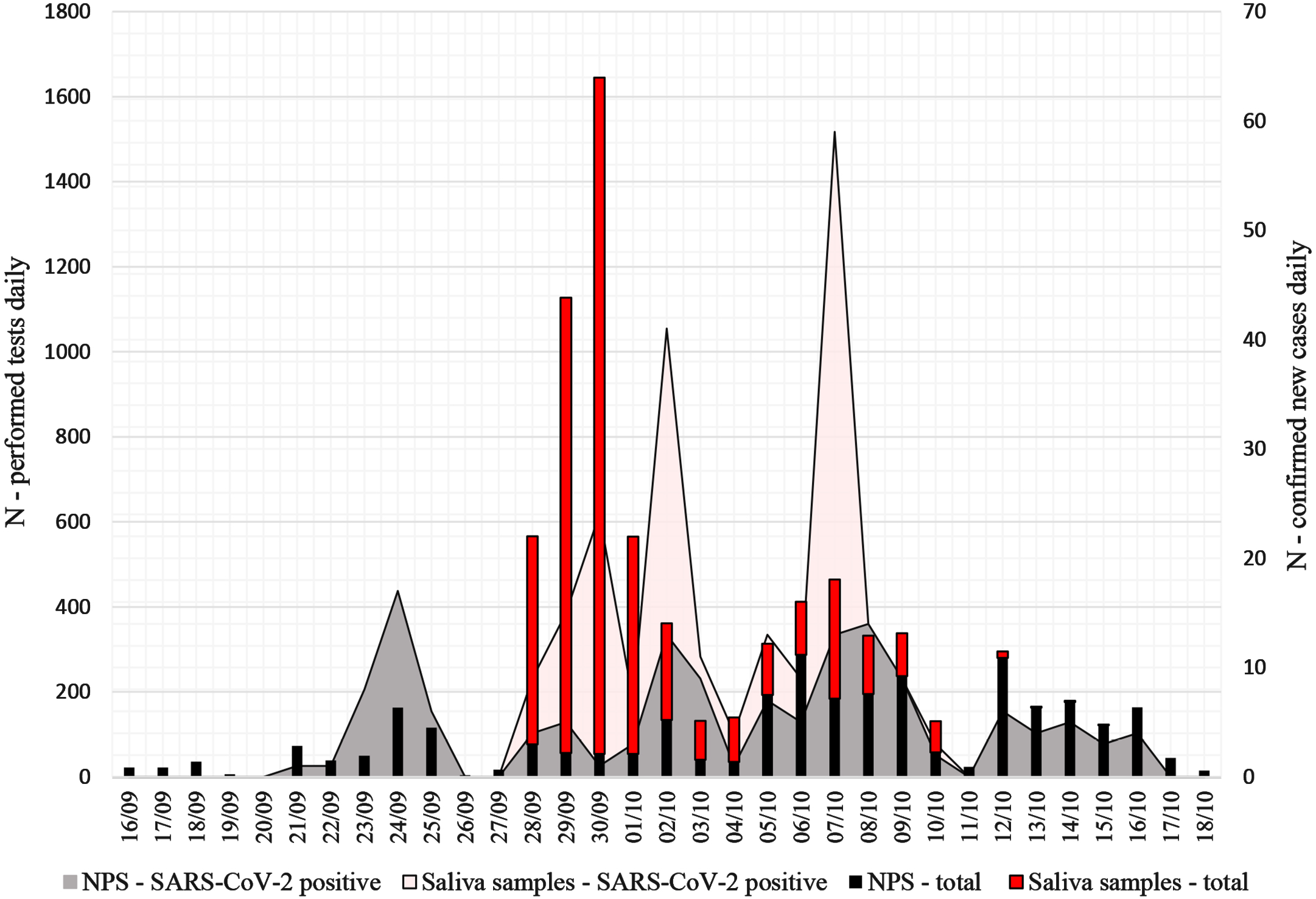

## Discussion

The findings of our study can be summarized as follows: (a) for the detection of SARS-CoV-2, saliva is a stable specimen with acceptable specificity and sensitivity at the early stages of infection; (b) saliva specimen is appropriate for pooling, with accurate diagnostic performance.

Saliva has been identified as a reliable testing specimen for SARS-CoV-2 using the RT-PCR approach in several recent studies (7–9). In the updated European Centre for Disease Prevention and Control (ECDC) recommendations, saliva is also mentioned as a convenient specimen for SARS-CoV-2 testing (10,11). The FDA has approved methods for SARS-CoV-2 testing using saliva in several laboratories (12,13). Nevertheless, saliva has been included in IFCC COVID-19 Guidelines on Molecular, Serological, and Biochemical/Haematological Testing as a promising sample type (14).

The high expression of ACE2 receptors in salivary gland cells is the main factor for virus affinity that could lead to active replication and transmission by saliva droplets expelled into the air during coughing or sneezing (15,16).

It is essential to establish standardized sample collection procedures, safe logistics, and reliable testing methods that meet performance requirements. Our stability testing confirmed that saliva samples stayed equally positive up to 24h.

In the analytical context, saliva can have the same or even better performance than NPS (17). It has been previously confirmed to be highly sensitive and specific at the early stage of infection 0-14 days after onset of symptoms and in asymptomatic cases (17,18). Therefore, it is essential to take into consideration the relationship between the dynamics of viral load, Ct values, and the number of days after the onset of symptoms. As described in prior publications, the Ct value is important in determining the infectiousness of a patient sample. Ct values above 34 do not emit infectious virus particles, and values between 27-34 show low viability of the viral load. Samples with a Ct value of 13-17 show positive virus viability (19). Ct values may vary depending on the different assays by up to 5 cycles for the same sample (20). Variation also appears due to the quality of the specimen obtained and the different treatment methods used to prepare samples for testing (21). As expected, we got some variation between the NPS and saliva sample types. In our pilot testing using 44 positive saliva samples we found slightly higher (13%) Ct values in pools, compared to individual tests. Similar findings have been published by other researchers (22). This observed slight difference still allows for the safe use of saliva pooling for surveillance purposes with sufficient diagnostic accuracy. Nevertheless, pooling strategies are described in the latest update of COVID-19 testing strategies and objectives published by the ECDC (11). There is evidence of benefits from pooling in low prevalence countries with a low proportion of positive samples – up to 5%. Recent publications indicate that for populations with a prevalence of less than 1% the testing of saliva pools of 10 or 20 samples is more beneficial, and our independent calculations also bear this out. Comparing the correlation between different pool sizes (usefulness and efficiency) with test positivity rate, we concluded that a pool size of 10 is more efficient than a pool size of 5. The calculations are based on test price, reimbursement conditions, and the number of tests per sample (when using a pooling strategy) (23,24).

The pooling strategy for self-collected saliva samples is the optimal solution that saves resources and reduces the testing time (25). In our real-life field test on a “perifocal” population, we successfully tested nearly a third of the citizens of the town of Kuldiga in three days. This allowed for fast identification of asymptomatic SARS-CoV-2 cases and those with mild symptoms to enable timely contact tracing and outbreak containment. The use of nasopharyngeal swabs in such a situation would have required more staff and time with possible patient compliance issues.

Additionally, improvements may reasonably be expected in terms of organizing the distribution of self-sampling kits to patients as well as the logistics of returning the kits to the laboratory that would further increase the usefulness of screening using a pooled testing strategy.

There were some limitations in our study: the size of the cohort of participants was limited, the selected laboratory-developed RT-PCR method for evaluation was used, also a lack of multicenter observation. Future studies extending the cohort are needed to confirm current findings and provide the implications in clinical practice.

## Conclusions

Our findings confirm the stability of SARS-CoV-2 RNA in saliva, showing acceptable performance in terms of specificity and sensitivity at the early stages of infection, and the advantages of testing using a pooling strategy in a low prevalence population. Ct values and detected/missing gene information are favorable for the interpretation of the results from several aspects, e.g. epidemiological investigation, determination if the patient is infectious at the current stage, etc. Self-sampling makes the procedure faster, safer, and requires fewer resources. The targeted distribution of test kits among a population with a known outbreak significantly increased the positivity rate. Saliva pool testing on a large scale provides an additional tool to take timely measures and contain outbreaks.

## Data Availability

All data generated or analysed during this study are included in this published article.

## Ethical aspects

The study was approved by the Ethics Committee of the Pauls Stradins Clinical University Hospital (record No 300720-18L).

## Acknowledgments

We gratefully acknowledge all participants for their commitment to participate and their time. Special thanks to all trial team members in the E.Gulbja Laboratory for their dedication and work which made this study possible and Dr. A.Gramatniece for editorial help.

## Author contributions

Gavars D - the conception and design of the study, Gavars M, Perminovs D, and Metla Z – validation of the method and interpretation of the data, Stasulans J, Stana J - patient recruitment and data collection, drafting of the article, Tauckels E - data analysis, Gulbis E, and Dumpis U – revising the article critically, final approval of the version to be submitted.

## Transparency declaration

All authors declare that there are no conflicts of interest.

## Funding institution

Ministry of Education and Science, Republic of Latvia. Funding number: VPP-COVID-2020/1-0008.

Additional funding was provided by E.Gulbja laboratory.

## Notes

### Competing Interest Statement

The authors have declared no competing interest.

### Author Declarations

The study was approved by Ethics Committee of Pauls Stradins Clinical university hospital (record No 300720-18L).

## References

1. Rapid Risk Assessment: Coronavirus disease 2019 (COVID-19) in the EU/EEA and the UK– ninth update [Internet]. European Centre for Disease Prevention and Control. 2020 [cited 2020 Nov 16]. Available from: https://www.ecdc.europa.eu/en/publications-data/rapid-risk-assessment-coronavirus-disease-2019-covid-19-pandemic-ninth-update

2. Laboratory testing for 2019 novel coronavirus (2019-nCoV) in suspected human cases [Internet]. [cited 2020 Nov 16]. Available from: https://www.who.int/publications-detail-redirect/10665-331501

3. COVID-19: Overcoming supply shortages for diagnostic testing | McKinsey [Internet]. [cited 2020 Nov 16]. Available from: https://www.mckinsey.com/industries/pharmaceuticals-and-medical-products/our-insights/covid-19-overcoming-supply-shortages-for-diagnostic-testing#

4. Jeong HW, Kim S-M, Kim H-S, Kim Y-I, Kim JH, Cho JY, et al. Viable SARS-CoV-2 in various specimens from COVID-19 patients. Clin Microbiol Infect. 2020 Nov;26(11):1520–4.

5. COVID-19 situation update for the EU/EEA and the UK, as of 16 November 2020 [Internet]. European Centre for Disease Prevention and Control. [cited 2020 Nov 16]. Available from: https://www.ecdc.europa.eu/en/cases-2019-ncov-eueea

6. Marty FM, Chen K, Verrill KA. How to Obtain a Nasopharyngeal Swab Specimen. N Engl J Med. 2020 May 28;382(22):e76.

7. Fakheran O, Dehghannejad M, Khademi A. Saliva as a diagnostic specimen for detection of SARS-CoV-2 in suspected patients: a scoping review. Infect Dis Poverty. 2020 Jul 22;9(1):100.

8. Hamid H, Khurshid Z, Adanir N, Zafar MS, Zohaib S. COVID-19 Pandemic and Role of Human Saliva as a Testing Biofluid in Point-of-Care Technology. Eur J Dent [Internet]. 2020 Jun 3 [cited 2020 Nov 16]; Available from: http://www.thieme-connect.de/DOI/DOI?10.1055/s-0040-1713020

9. Czumbel LM, Kiss S, Farkas N, Mandel I, Hegyi A, Nagy Á, et al. Saliva as a Candidate for COVID-19 Diagnostic Testing: A Meta-Analysis. Front Med. 2020 Aug 4;7:465–465.

10. Objectives for COVID-19 testing in school settings - first update, 21 August 2020. Tech Rep. 2020;4.

11. COVID-19 testing strategies and objectives [Internet]. European Centre for Disease Prevention and Control. 2020 [cited 2020 Nov 16]. Available from: https://www.ecdc.europa.eu/en/publications-data/covid-19-testing-strategies-and-objectives

12. Commissioner O of the. Coronavirus (COVID-19) Update: FDA Authorizes First Diagnostic Test Using At-Home Collection of Saliva Specimens [Internet]. FDA. FDA; 2020 [cited 2020 Nov 16]. Available from: https://www.fda.gov/news-events/press-announcements/coronavirus-covid-19-update-fda-authorizes-first-diagnostic-test-using-home-collection-saliva

13. Rutgers Launches Genetic Testing Service for New Coronavirus | Rutgers University [Internet]. [cited 2020 Nov 16]. Available from: https://www.rutgers.edu/news/rutgers-launches-genetic-testing-service-new-coronavirus

14. Bohn M, Mancini N, Loh P, Wang C-B, Grimmler M, Gramegna M, et al. IFCC interim guidelines on molecular testing of SARS-CoV-2 infection. Clin Chem Lab Med. 2020 Oct 7;

15. Liu L, Wei Q, Alvarez X, Wang H, Du Y, Zhu H, et al. Epithelial Cells Lining Salivary Gland Ducts Are Early Target Cells of Severe Acute Respiratory Syndrome Coronavirus Infection in the Upper Respiratory Tracts of Rhesus Macaques. J Virol. 2011 Apr 15;85(8):4025–30.

16. Xu J, Li Y, Gan F, Du Y, Yao Y. Salivary Glands: Potential Reservoirs for COVID-19 Asymptomatic Infection. J Dent Res. 2020;99(8):989.

17. Wyllie AL, Fournier J, Casanovas-Massana A, Campbell M, Tokuyama M, Vijayakumar P, et al. Saliva or Nasopharyngeal Swab Specimens for Detection of SARS-CoV-2. N Engl J Med. 2020;4.

18. Zhang W, Du R-H, Li B, Zheng X-S, Yang X-L, Hu B, et al. Molecular and serological investigation of 2019-nCoV infected patients: implication of multiple shedding routes. Emerg Microbes Infect. 2020 Feb 17;9(1):386–9.

19. La Scola B, Le Bideau M, Andreani J, Hoang VT, Grimaldier C, Colson P, et al. Viral RNA load as determined by cell culture as a management tool for discharge of SARS-CoV-2 patients from infectious disease wards. Eur J Clin Microbiol Infect Dis. 2020 Apr 27;1–3.

20. Nalla AK, Casto AM, Huang M-LW, Perchetti GA, Sampoleo R, Shrestha L, et al. Comparative Performance of SARS-CoV-2 Detection Assays Using Seven Different Primer-Probe Sets and One Assay Kit. J Clin Microbiol. 2020 26;58(6).

21. Rodino KG, Espy MJ, Buckwalter SP, Walchak RC, Germer JJ, Fernholz E, et al. Evaluation of Saline, Phosphate-Buffered Saline, and Minimum Essential Medium as Potential Alternatives to Viral Transport Media for SARS-CoV-2 Testing. McAdam AJ, editor. J Clin Microbiol. 2020 May 26;58(6):e00590–20.

22. Lohse S, Pfuhl T, Berkó-Göttel B, Rissland J, Geißler T, Gärtner B, et al. Pooling of samples for testing for SARS-CoV-2 in asymptomatic people. Lancet Infect Dis. 2020 Nov 1;20(11):1231–2.

23. Regen F, Eren N, Heuser I, Hellmann-Regen J. A simple approach to optimum pool size for pooled SARS-CoV-2 testing. Int J Infect Dis. 2020 Nov 1;100:324–6.

24. Mutesa L, Ndishimye P, Butera Y, Souopgui J, Uwineza A, Rutayisire R, et al. A pooled testing strategy for identifying SARS-CoV-2 at low prevalence. Nature. 2020 Oct 21;1–8.

25. Fogarty A, Joseph A, Shaw D. Pooled saliva samples for COVID-19 surveillance programme. Lancet Respir Med. 2020;8(11):1078–80.

